# Comparing effect estimates in randomized trials and observational studies from the same population: an application to percutaneous coronary intervention

**DOI:** 10.1101/2021.02.01.21250739

**Authors:** Anthony A Matthews, Karolina Szummer, Issa J Dahabreh, Bertil Lindahl, David Erlinge, Maria Feychting, Tomas Jernberg, Anita Berglund, Miguel A Hernán

**Author notes:** Correspondence* Anthony A Matthews, Address: Unit of Epidemiology, Institute of Environmental Medicine, Karolinska Institutet, Stockholm, Sweden.

## Abstract

**Background:** The ability for real world data to deliver similar results as a trial that asks the same question about the risks or benefits of a clinical intervention can be restricted not only by lack of randomization, but also limited information on eligibility criteria and outcomes. To understand when results from observational studies and randomized trials are comparable, we carried out an observational emulation of a target trial designed to ask similar questions as the VALIDATE randomized trial. VALIDATE compared the effect of bivalirudin and heparin during percutaneous coronary intervention on the risk of death, myocardial infarction, and bleeding across Sweden.

**Methods:** We specified the protocol of a target trial similar to the VALIDATE trial protocol, then emulated the target trial in the period before the trial took place using data from the SWEDEHEART registry; the same registry in which the trial was undertaken.

**Results:** The target trial emulation and the VALIDATE trial both estimated no difference in the effect of bivalirudin and heparin on the risk of death or myocardial infarction by 180 days: emulation risk ratio for death 1.21 (0.88, 1.54); VALIDATE hazard ratio for death 1.05 (0.78, 1.41). The observational data, however, could not capture less severe cases of bleeding, resulting in an inability to define a bleeding outcome like the trial, and could not account for intractable confounding early in follow-up (risk ratio for death by 14 days 1.85 (0.95, 3.63)).

**Conclusion:** Using real world data to emulate a target trial can deliver accurate long-term effect estimates. Yet, even with rich observational data, it is not always possible to estimate the short-term effect of interventions, or the effect on outcomes for which data are not routinely collected. If registries included information on reasons for treatment decisions, researchers may be better positioned to identify important confounders.

## Background

Observational analyses of routinely collected healthcare data, a form of “real world evidence”, are often used to evaluate the benefits and risks of clinical interventions for acute coronary syndromes. Most criticisms of these observational analyses revolve around the lack of randomized assignment of the treatment strategies under comparison, which may result in confounded effect estimates.^1-3^ Despite this limitation, routinely collected healthcare data can be used to explicitly emulate target trials, as recent applications in diverse clinical areas have shown.^4-8^

Routinely collected observational data can only be used to emulate pragmatic trials, but not trials with placebo control and blind treatment assignment, or those that rely on detailed data collection to determine eligibility or ascertain outcomes.^9^ Specifically, the lack of detailed information on eligibility criteria and outcomes restricts the type of causal questions that can be answered using real world data, a point that is not always emphasized in discussions about the topic.^4^

To study the strengths and limitations of observational analyses that emulate a trial, a near ideal scenario is to compare a registry-based randomized trial versus the emulation of a target trial with the same protocol as the registry-based randomized trial using observational data from the same registry.^10^ This comparison ensures that the causal question is asked in the same population and health care setting.^11^ The Swedish Web System for Enhancement and Development of Evidence-based Care in Heart Disease Evaluated According to Recommended Therapies (SWEDEHEART) is a national quality registry of myocardial infarction, coronary intervention, and heart surgery, which includes longitudinal information on demographic and clinical characteristics, use of therapeutic and preventive services, diagnostic procedures, and various measures of health care utilization for the whole of Sweden.^12^ By using the registry to enroll participants, randomize interventions, and report outcomes, SWEDEHEART can be used to run randomized trials nested within the registry.^13^ An example of a registry-based randomized trial within SWEDEHEART is the VALIDATE trial, which compared two anticoagulation interventions, bivalirudin and heparin, over 180 days in patients with acute myocardial infarction undergoing percutaneous coronary intervention in Sweden.^14^

Here, we use the observational data from the SWEDEHEART registry to emulate a target trial designed to answer similar questions as the VALIDATE trial, and we outline conditions under which one can successfully design a target trial and carry out an observational emulation of this target trial. We describe the VALIDATE randomized trial and the observational data from the SWEDEHEART registry, the specification of the target trial, and the emulation procedures. Our example illustrates the opportunities and limitations of healthcare registries to emulate a target trial.

## VALIDATE: the index randomized trial

### Trial design and analysis

VALIDATE was a multicenter, randomized, controlled, open-label clinical trial carried out between June 2014 and September 2016. Individuals in the SWEDEHEART registry were eligible to participate if they were admitted to hospital with a diagnosis of ST-segment elevated myocardial infarction (STEMI) or non-ST-segment elevated myocardial infarction (NSTEMI), urgent percutaneous coronary intervention was planned in one of 25 of the 29 percutaneous coronary intervention centers in Sweden, and some other eligibility criteria were met (see Table 1). Individuals who accepted the invitation to participate were randomly assigned to receive either intravenous bivalirudin (0.75 mg per kg) or intraarterial unfractionated heparin (5,000 U/ml) under percutaneous coronary intervention. The primary end point was the composite of death from any cause, myocardial infarction, or major bleeding by 180 days. The individual components of the composite event were also assessed. Research nurses screened for clinical end-point events by contacting the patients or first-degree relatives by telephone 7 days and 180 days after percutaneous coronary intervention. If the patient or relatives could not be contacted after the nurses had placed repeated telephone calls and mailed a letter, information was collected through review of hospital records. The intention-to-treat analysis, described in detail elsewhere, relied on the comparison of 180-day risk differences and hazard ratios.^14^

**Table 1.** Description of VALIDATE randomized trial, target trial, and target trial emulation using SWEDEHEART register.

| Protocol Component | VALIDATE trial | Target trial | Target trial emulation using SWEDEHEART |
| --- | --- | --- | --- |
| **Eligibility criteria** | - Age 18 years or older between June 2014 and September 2016<br>- Diagnosis of non-ST-elevation myocardial infarction as defined by guidelines (positive troponin) or ST-elevation myocardial infarction as defined by chest pain for at least 30 minutes and an ECG with new ST-segment elevation in two or more contiguous leads of $\geq 0.2$ mV in leads V2-V3 and/or $\geq 0.1$ mV in other leads or a probable new-onset left bundle branch block<br>- Therapeutic percutaneous coronary intervention (not primarily diagnostic percutaneous coronary intervention) after angiography in one of 25 (of 29) selected clinics<br>- Ability to provide informed consent: In patients with ST-elevation myocardial infarction, witnessed oral consent was obtained after angiography and before randomization. Within the following 24 hours, after written information about the trial had been provided, the patients confirmed further participation by providing written informed consent. In patients with non-ST-elevation myocardial infarction, written consent was obtained before angiography.<br>- Treated with bolus dose of ticagrelor, prasugrel, or cangrelor before start of percutaneous coronary intervention<br>- Treated with aspirin in accordance with local practices<br>- Life expectancy greater than one year<br>- Patients with known ongoing bleeding<br>- No heparin >5000U before arriving to percutaneous coronary intervention lab. Up to 3000U of heparin allowed during angiography.<br>- No uncontrolled hypertension, subacute bacterial endocarditis, severe renal (GFR < 30 ml/min) and /or liver dysfunction, thrombocytopenia or thrombocyte function defects<br>- No contraindication for the study medications<br>- No GpIIb/IIIa-inhibitors given or are pre-planned to be given during the procedure | Same as VALIDATE apart from:<br>- Study period between January 2012 and May 2014<br>- Without asking for informed consent<br>- Known terminal disease defined as palliative care, dialysis, dementia, or aggressive cancer (pancreatic, mesothelioma, lung, liver, stomach, or brain) in 3 years prior to percutaneous coronary intervention<br>- Unable to identify ongoing bleeding<br>- No information on dose of heparin prior to percutaneous coronary intervention, so excluded all individuals given any prior heparin<br>- Unable to identify contraindications to study medications<br>- Individuals that died on the day of percutaneous coronary intervention were excluded and identification of outcomes start from day after percutaneous coronary intervention | Same as target trial |
| **Treatment strategies** | (1) Bivalirudin: intravenous bolus of 0.75 mg per kilogram of body weight followed by an infusion of 1.75 mg per kilogram per hour. Treatment was started as soon as percutaneous coronary intervention of the culprit lesion was planned. Continuation of the bivalirudin infusion after percutaneous coronary intervention until completion of the last vial is strongly recommended.<br>(2) Heparin: 70 to 100U per kilogram. | (1) Individual administered bivalirudin under percutaneous coronary intervention<br>(2) Individual administered heparin under percutaneous coronary intervention<br>- If patient given both bivalirudin and heparin, then it was assumed that heparin was low dose intraarterial administration of up to 3000U, and treatment strategy was therefore bivalirudin, as no information was available on dosage. | Same as target trial. Individuals were classified according to the strategy that their data were compatible with at date of percutaneous coronary intervention, |
| **Treatment assignment** | Before percutaneous coronary intervention, patients were randomly assigned to receive in an open-label fashion either intravenous bivalirudin or intraarterial unfractionated heparin. Randomization was performed in a 1:1 ratio with stratification according to type of myocardial infarction (ST-elevation myocardial infarction or non-ST-elevation myocardial infarction) and hospital. | - Individuals randomized to a treatment strategy under percutaneous coronary intervention and were aware of the assigned strategy. | Assignment was treated as if randomized within levels of the following baseline covariates: severity of myocardial infarction (STEMI/NSTEMI, Killip class), center where percutaneous coronary intervention took place (university hospital/non-university hospital), demographics (sex, age), lifestyle characteristics (weight), laboratory measurements (GFR), diagnoses (bleeding, anemia), and medications (warfarin, novel oral anti-coagulants (NOAC), P2Y12 inhibitors) |
| **Outcomes** | - Composite of death from any cause, myocardial infarction, or major bleeding events by 180 days.<br>- Each separate component of the composite. | Same as VALIDATE, but outcomes identified as follows:<br>- Composite death from any cause identified from the Swedish Cause of Death register, myocardial infarction identified from the SWEDEHEART register, and major bleeding events identified from the Swedish Inpatient and Outpatient registers or the SWEDEHEART register by 180 days | Same as target trial |
| **Follow-up** | Starts at treatment assignment and ends at date of first outcome (separately for analysis of each outcome), migration, or 180 days after baseline. | Same as VALIDATE apart from:<br>- Unable to identify migration date | Same as target trial |
| **Causal contrast** | Intention to treat effect | Intention to treat effect | Observational analogue of the intention-to-treat effect |
| **Statistical analysis** | - Kaplan-Meier plots<br>- Treatment differences were assessed with the use of the log-rank test and Cox regression. | Same as VALIDATE apart from:<br>- Kaplan-Meier plots adjusted for baseline covariates via inverse probability weighting<br>- Treatment differences were assessed with use of pooled logistic regression models with a flexible time-varying intercept and product terms between the treatment and time. Baseline covariates adjusted for using two alternative approaches: inverse probability weighting (the baseline covariates were not included in the outcome model) and standardization (the baseline covariates were included in the outcome model). | Same as target trial |

### Trial results

A total of 6,006 patients underwent randomization in the VALIDATE trial, 3,004 were assigned to bivalirudin and 3,002 were assigned to heparin. The risk of the composite end point did not differ between the treatment groups 30 days after percutaneous coronary intervention. By 180 days, the composite endpoint occurred in 12.3% of patients (369 of 3004) in the bivalirudin group and in 12.8% (383 of 3002) in the heparin group, with a hazard ratio of 0.96 (95% CI: 0.83, 1.10). Death occurred in 2.9% of patients in the bivalirudin group and in 2.8% in the heparin group (without accounting for censoring), with a hazard ratio of 1.05 (95% CI: 0.78, 1.41). Myocardial infarction occurred in 2.0% of patients in the bivalirudin group and in 2.4% in the heparin group, with a hazard ratio of 0.84 (95% CI: 0.60, 1.19). Major bleeding occurred in 8.6% of patients in both the bivalirudin and heparin groups, with a hazard ratio of 1.00 (95% CI: 0.84, 1.19).

## Specifying and emulating a target trial in the SWEDEHEART registry

### Data sources

SWEDEHEART collects data from all patients hospitalized for acute coronary syndrome or undergoing coronary or valvular intervention for any indication in all relevant hospitals across Sweden.^13^ The registry was officially launched in 2009 when four existing cardiovascular healthcare quality registries were merged: the Register of Information and Knowledge About Swedish Heart Intensive Care Admissions (RIKSHIA), the Swedish Coronary Angiography and Angioplasty Registry (SCAAR), the Swedish Heart Surgery Register and the National Registry of Secondary Prevention (SEPHIA), and the Swedish Heart Surgery Registry. SWEDEHEART was used to collect information for patients when they were randomized in the VALIDATE trial, hence the target trial was emulated in the same population as the original trial and the data collection process was broadly similar between the two studies. SWEDEHEART is also linked to the Inpatient and Outpatient Register, which records all primary and secondary diagnoses and procedures from inpatient hospitalizations and outpatient specialist care visits across Sweden; the Swedish Cause of Death register, which records all deaths and causes of death; and the Prescribed Drug register, which collects information on all dispensed medications.^15-17^

The approach for emulating a target trial similar to the VALIDATE trial had two steps: 1) specifying the protocol of the target trial, and 2) emulating the target trial using the observational data from the SWEDEHEART registry. Our target trial has the same protocol as the VALIDATE trial, with exceptions only when the observational data were not adequate to identify the information collected in the trial. Table 1 summarizes the target trial protocol and outlines the emulation procedure described below.

### Eligibility criteria

We identified individuals in the SWEDEHEART registry data that met the target trial’s eligibility criteria. The eligibility criteria for the target trial were the same as the VALIDATE trial with six exceptions. First, the study period was between January 2012 to May 2014, which immediately precedes the period of the VALIDATE trial, as all eligible patients during that period were considered in the VALIDATE trial. Second, no informed consent was asked and hence the target trial could not exclude individuals who would have no been asked or declined participation if asked. Third, the target trial used a proxy measure to indicate known terminal disease with life expectancy less than a year (palliative care; dialysis; dementia; or malignant disease, including pancreatic, lung, liver, stomach, and brain cancer, or mesothelioma). Fourth, some eligibility criteria could not be applied in the target trial due to unavailable data (ongoing bleeding, contraindications to study medication). Fifth, there was no information on dose of heparin prior to percutaneous coronary intervention so we excluded all individuals with any prior heparin. Sixth, individuals that died on the day of percutaneous coronary intervention were excluded and identification of outcomes started from the day after percutaneous coronary intervention as it was not possible to distinguish if outcome events such as myocardial infarction and bleeding occurred before or after percutaneous coronary intervention when the events occurred on the same day as the procedure.

### Treatment strategies

The treatment strategies in the target trial closely mimicked those in the VALIDATE trial and were 1) administration of bivalirudin or 2) administration of heparin during percutaneous coronary intervention. The SWEDHEART registry contains information on the anticoagulant given under percutaneous coronary intervention, but does not contain information on dosage. In the target trial, we therefore assumed that if a patient was given both treatments, then the heparin was low dose, and the treatment strategy was defined as bivalirudin.

### Treatment assignment

We classified eligible individuals in the SWEDEHEART registry into two groups according to the strategy their data were compatible with at baseline. That is, our emulation presumes that all individuals assigned to a treatment strategy ended up receiving it. Our emulation treats individuals as if they were randomly assigned to a treatment strategy conditional on the baseline covariates: severity of myocardial infarction (STEMI/NSTEMI, Killip class, angiography finding), center where percutaneous coronary intervention took place (university hospital/non-university hospital), demographics (sex, age), lifestyle characteristics (weight, smoking), laboratory measurements (GFR, heart rate, systolic blood pressure, diastolic blood pressure), diagnoses (diabetes, severe bleeding, anemia), medications (warfarin, anti-hypertensives, lipid lowering treatment), and prior cardiovascular disease or cardiovascular procedures (myocardial infarction, percutaneous coronary intervention, coronary artery bypass grafting). See Appendix 1 for further details on covariates and their definitions.

### Outcomes

The outcomes in the target trial were the same as the VALIDATE trial and included death from any cause, myocardial infarction, major bleeding events, and a composite of all above outcomes. Death was identified from the Cause of Death register, myocardial infarction from the SWEDEHEART registry, and bleeding from the Inpatient and Outpatient register or SWEDEHEART registry (full outcome definitions in Appendix 2).

### Follow-up

Each individual was followed from the day following percutaneous coronary intervention until the outcome of interest or 180 days, whichever occurred first. In the myocardial infarction and bleeding analyses, individuals were not censored if they died during follow-up, which is valid when estimating the total effect of treatment, but we undertook a sensitivity analysis to explore censoring at death (Sensitivity 10).^18^ Outcome data on those who migrated out of Sweden were unavailable, but the probability of migration during such a short period is low.

### Causal contrast

In the target trial, the intention-to-treat effect is the effect of being assigned to bivalirudin versus heparin at baseline on the risk of death, myocardial infarction, bleeding, and a composite outcome of all outcomes. Our emulation focuses on an observational analog of the intention-to-treat effect under the assumption that all individuals received the treatment they were assigned to.

### Statistical analysis

We estimated the 14, 30, and 180-day risk (cumulative incidence), risk difference, and risk ratio for each outcome using pooled logistic regression models with a flexible time-varying intercept and product terms between the treatment and time.^19^ We adjusted for baseline covariates using two alternative approaches: inverse probability weighting and standardization. We truncated the stabilized inverse probability weights at their 99^th^ percentile to prevent outliers with extreme weights from influencing effect estimates. We plotted unweighted and inverse probability weighted Kaplan-Meier survival curves for all outcomes. We used nonparametric bootstrapping with 500 samples to calculate all 95% confidence intervals (CI). Further details on our modelling approach are in Appendix 3.

We repeated the 180-day analysis using inverse probability weighted models stratified by myocardial infarction type (STEMI/NSTEMI) to explore effect modification. Finally, we carried out several sensitivity analyses using inverse probability weighted models to assess the sensitivity of the eligibility criteria, treatment strategies, outcomes, confounders, follow-up, and analysis. Full details on sensitivity analyses are in Appendix 4. All analysis code is available at: https://github.com/tonymatthews/validate.

## Results

Figure 1 shows a flowchart of patient selection for the target trial emulation. Table 2 shows the baseline characteristics of the 4,940 eligible patients, of whom 2,634 were given bivalirudin and 2,306 heparin. Compared with those in the heparin group, patients in the bivalirudin group were more likely to be diagnosed with STEMI over NSTEMI before percutaneous coronary intervention (81% vs 17%).

**Table 2.**
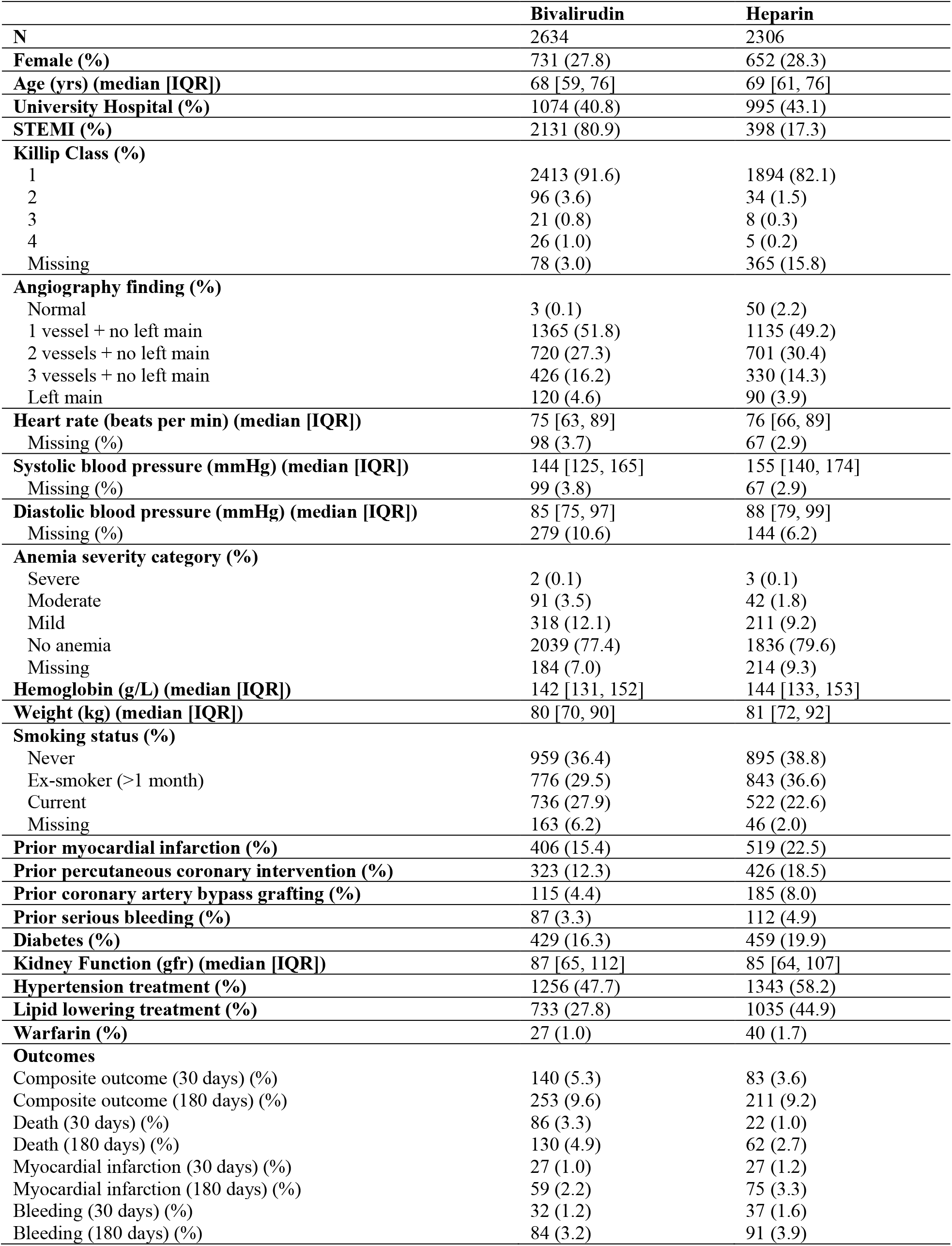
Baseline characteristics of eligible individuals for the observational emulation of a target trial of bivalirudin vs. heparin, SWEDEHEART Register, 2012-2014.

|  | **Bivalirudin** | **Heparin** |
| --- | --- | --- |
| **N** | 2634 | 2306 |
| **Female (%)** | 731 (27.8) | 652 (28.3) |
| **Age (yrs) (median [IQR])** | 68 [59, 76] | 69 [61, 76] |
| **University Hospital (%)** | 1074 (40.8) | 995 (43.1) |
| **STEMI (%)** | 2131 (80.9) | 398 (17.3) |
| **Killip Class (%)** |  |  |
| 1 | 2413 (91.6) | 1894 (82.1) |
| 2 | 96 (3.6) | 34 (1.5) |
| 3 | 21 (0.8) | 8 (0.3) |
| 4 | 26 (1.0) | 5 (0.2) |
| Missing | 78 (3.0) | 365 (15.8) |
| **Angiography finding (%)** |  |  |
| Normal | 3 (0.1) | 50 (2.2) |
| 1 vessel + no left main | 1365 (51.8) | 1135 (49.2) |
| 2 vessels + no left main | 720 (27.3) | 701 (30.4) |
| 3 vessels + no left main | 426 (16.2) | 330 (14.3) |
| Left main | 120 (4.6) | 90 (3.9) |
| **Heart rate (beats per min) (median [IQR])** | 75 [63, 89] | 76 [66, 89] |
| Missing (%) | 98 (3.7) | 67 (2.9) |
| **Systolic blood pressure (mmHg) (median [IQR])** | 144 [125, 165] | 155 [140, 174] |
| Missing (%) | 99 (3.8) | 67 (2.9) |
| **Diastolic blood pressure (mmHg) (median [IQR])** | 85 [75, 97] | 88 [79, 99] |
| Missing (%) | 279 (10.6) | 144 (6.2) |
| **Anemia severity category (%)** |  |  |
| Severe | 2 (0.1) | 3 (0.1) |
| Moderate | 91 (3.5) | 42 (1.8) |
| Mild | 318 (12.1) | 211 (9.2) |
| No anemia | 2039 (77.4) | 1836 (79.6) |
| Missing | 184 (7.0) | 214 (9.3) |
| **Hemoglobin (g/L) (median [IQR])** | 142 [131, 152] | 144 [133, 153] |
| **Weight (kg) (median [IQR])** | 80 [70, 90] | 81 [72, 92] |
| **Smoking status (%)** |  |  |
| Never | 959 (36.4) | 895 (38.8) |
| Ex-smoker (>1 month) | 776 (29.5) | 843 (36.6) |
| Current | 736 (27.9) | 522 (22.6) |
| Missing | 163 (6.2) | 46 (2.0) |
| **Prior myocardial infarction (%)** | 406 (15.4) | 519 (22.5) |
| **Prior percutaneous coronary intervention (%)** | 323 (12.3) | 426 (18.5) |
| **Prior coronary artery bypass grafting (%)** | 115 (4.4) | 185 (8.0) |
| **Prior serious bleeding (%)** | 87 (3.3) | 112 (4.9) |
| **Diabetes (%)** | 429 (16.3) | 459 (19.9) |
| **Kidney Function (gfr) (median [IQR])** | 87 [65, 112] | 85 [64, 107] |
| **Hypertension treatment (%)** | 1256 (47.7) | 1343 (58.2) |
| **Lipid lowering treatment (%)** | 733 (27.8) | 1035 (44.9) |
| **Warfarin (%)** | 27 (1.0) | 40 (1.7) |
| **Outcomes** |  |  |
| Composite outcome (30 days) (%) | 140 (5.3) | 83 (3.6) |
| Composite outcome (180 days) (%) | 253 (9.6) | 211 (9.2) |
| Death (30 days) (%) | 86 (3.3) | 22 (1.0) |
| Death (180 days) (%) | 130 (4.9) | 62 (2.7) |
| Myocardial infarction (30 days) (%) | 27 (1.0) | 27 (1.2) |
| Myocardial infarction (180 days) (%) | 59 (2.2) | 75 (3.3) |
| Bleeding (30 days) (%) | 32 (1.2) | 37 (1.6) |
| Bleeding (180 days) (%) | 84 (3.2) | 91 (3.9) |

**Figure 1.**
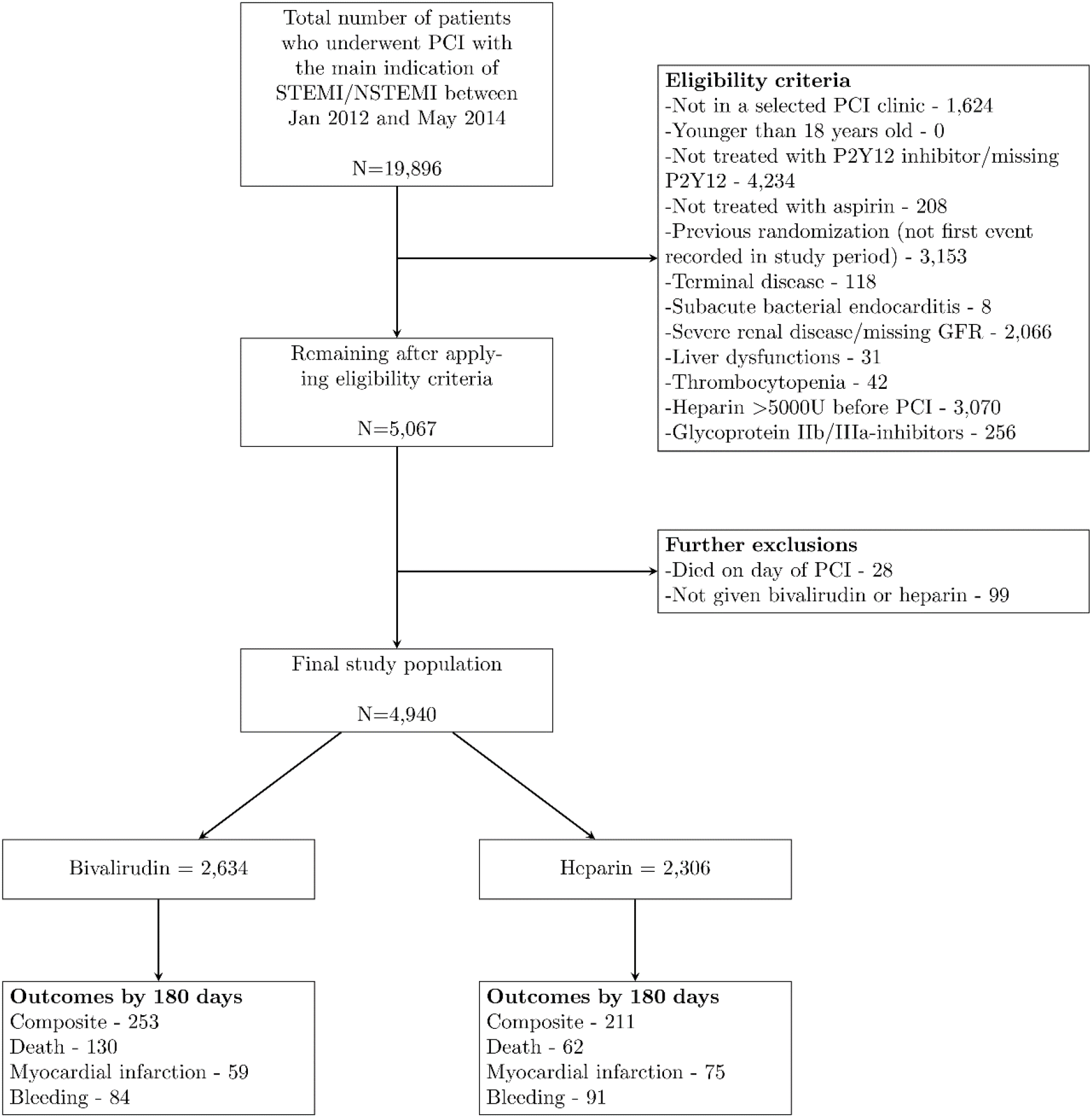
Flowchart of individuals eligible for the observational emulation of a target trial of bivalirudin vs. heparin, SWEDEHEART Register, 2012-2014.

Table 3 shows the estimated 180-day risks, risk differences, and risk ratios obtained via inverse probability weighting and standardization. The inverse probability weighted risk (95% CI) of the composite outcome was 9.3% (8.2%, 10.4%) in the bivalirudin group and 10.0% (8.7%, 11.3%) in the heparin group, which results in a risk difference of −0.7% (−2.5%, 1.1%) and a risk ratio of 0.93 (0.77, 1.12). The risk of death was 4.1% (3.4%, 4.8%) in the bivalirudin group and 3.4% (2.5%, 4.2%) in the heparin group, which results in a risk difference of 0.7% (−0.4%, 1.8%) and risk ratio of 1.21 (0.88, 1.68). The risk of myocardial infarction was 3.0% (2.3%, 3.8%) in the bivalirudin group and 2.8% (2.2%, 3.4%) in the heparin group, which results in a risk difference of 0.2% (−0.8%, 1.2%) and risk ratio of 1.08 (0.76, 1.54). The risk of bleeding was 3.2% (2.5%, 3.9%) in the bivalirudin group and 4.6% (3.7%, 5.6%) in the heparin group, which results in a risk difference of −1.4% (−2.6%, −0.3%) and risk ratio of 0.69 (0.50, 0.95). Standardized estimates were similar.

**Table 3.** Estimated 180-day risk, risk difference, and risk ratios from the observational emulation* of a target trial of bivalirudin vs. heparin, SWEDEHEART Register, 2012-2014.

| Outcome | Risk (%; 95% CI) |  | Risk difference<br>(%, 95% CI) | Risk ratio (95%<br>CI) |
| --- | --- | --- | --- | --- |
|  | Bivalirudin | Heparin |  |  |
| *Inverse-probability weighted* |  |  |  |  |
| Composite | 9.3 (8.2,10.4) | 10.0 (8.7,11.3) | -0.7 (-2.5,1.1) | 0.93 (0.77,1.12) |
| Death | 4.1 (3.4,4.8) | 3.4 (2.5,4.2) | 0.7 (-0.4,1.8) | 1.21 (0.88,1.68) |
| Myocardial infarction | 3.0 (2.3,3.8) | 2.8 (2.2,3.4) | 0.2 (-0.8,1.2) | 1.08 (0.76,1.54) |
| Bleeding | 3.2 (2.5,3.9) | 4.6 (3.7,5.6) | -1.4 (-2.6,-0.3) | 0.69 (0.50,0.95) |
| *Standardized* |  |  |  |  |
| Composite | 9.2 (7.5,10.8) | 9.9 (8.0,11.9) | -0.8 (-3.6,2.0) | 0.92 (0.74,1.14) |
| Death | 4.3 (3.2,5.4) | 3.4 (2.1,4.6) | 1.0 (-0.8,2.7) | 1.28 (0.92,1.80) |
| Myocardial infarction | 2.8 (1.6,3.9) | 2.7 (1.7,3.7) | 0.1 (-1.7,1.8) | 1.03 (0.66,1.59) |
| Bleeding | 3.0 (1.9,4.1) | 4.3 (2.6,5.9) | -1.3 (-3.6,1.1) | 0.70 (0.46,1.09) |

Figure 2 shows the inverse probability weighted survival curves (unweighted survival curves in Appendix 5). The myocardial infarction-free survival almost overlapped throughout the 180 days of follow-up. However, there was an elevated risk of death in the bivalirudin group compared with the heparin group in the 14 days after percutaneous coronary intervention. The short-term difference in risk of death can also be seen in the estimated risk difference and risk ratio by 14 and 30 days (Appendices 6 and 7).

**Figure 2.**
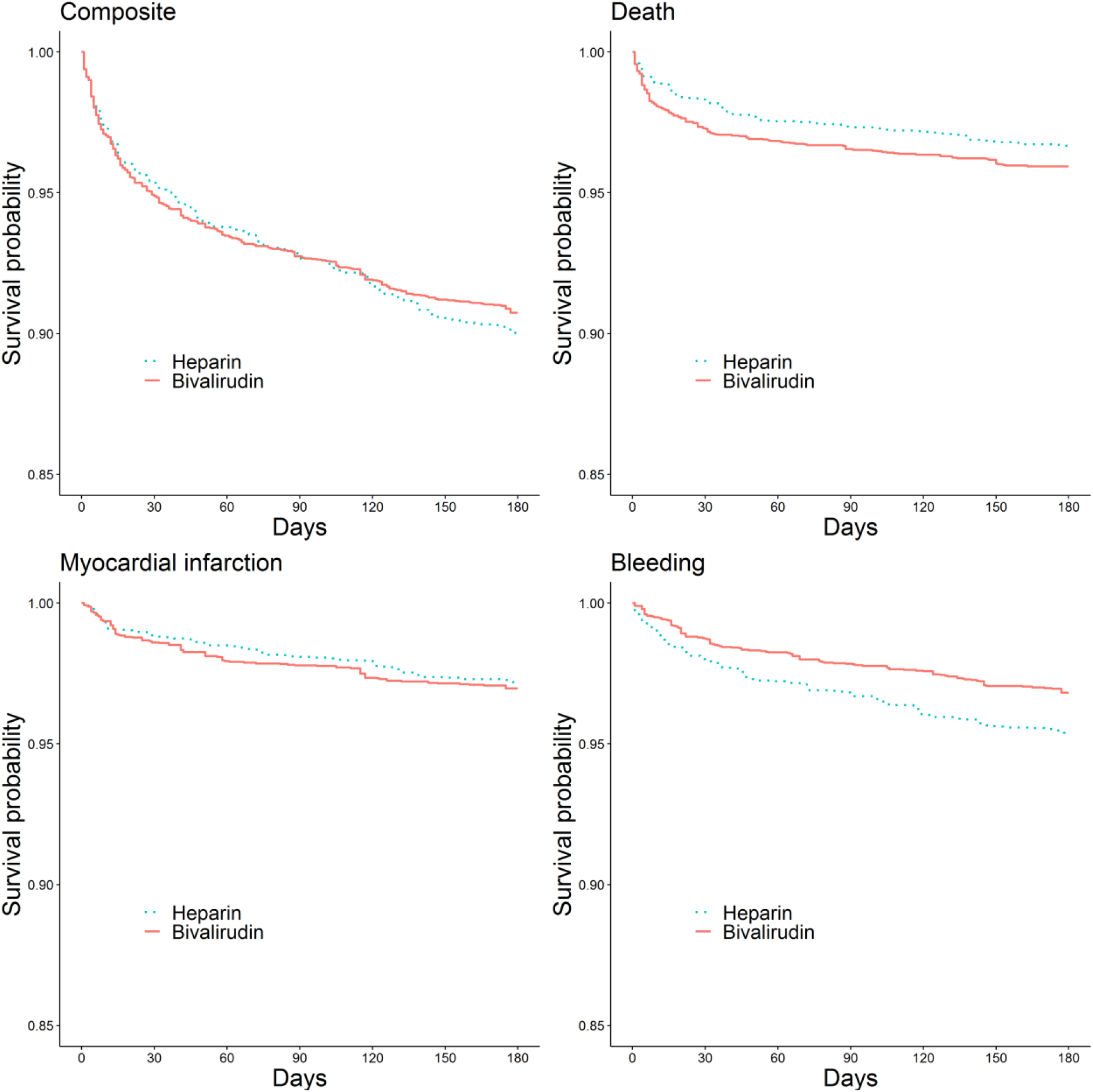
Inverse-probability weighted survival curves from an observational emulation of a target trial of bivalirudin vs. heparin, SWEDEHEART Register, 2012-2014*. *Adjusted at baseline for severity of myocardial infarction (STEMI/NSTEMI, Killip class, angiography finding), center where percutaneous coronary intervention took place (university hospital/non-university hospital), demographics (sex, age), lifestyle characteristics (weight, smoking), laboratory measurements (GFR, heart rate, systolic blood pressure, diastolic blood pressure), diagnoses (diabetes, severe bleeding, anemia), medications (warfarin, anti-hypertensives, lipid lowering treatment), and prior cardiovascular disease or cardiovascular procedures (myocardial infarction, percutaneous coronary intervention, coronary artery bypass grafting)

Appendix 8 shows the 180-day risks, risk differences, and risk ratios obtained via inverse probability weighting, stratified by myocardial infarction type, and there were no apparent differences between strata. Appendices 9-19 show results from sensitivity analyses; all results were broadly like those estimated in the main analyses.

## Discussion

We emulated a target trial similar to the VALIDATE trial using real world observational data from SWEDEHEART, the same register in which the trial was undertaken. There was broad agreement in the estimates for death and myocardial infarction from the randomized trial and the observational emulation of the target trial; both found little differences in risk between those given heparin and bivalirudin. However, there were some telling differences between the estimates from the VALIDATE trial and its observational emulation.

First, the target trial could not define a bleeding outcome similar to that in the VALIDATE trial, which was ascertained using a combination of phone calls to patients at 7 and 180 days after percutaneous coronary intervention and a review of hospital records from registers. Exclusively using the registers, as was done in the target trial, only allowed us to identify the most severe cases of bleeding. We also attempted to expand the bleeding outcome in a sensitivity analysis (outcome definition in Appendix 4 and results in Appendix 16), but we were still unable to fully capture less severe cases of bleeding. This difference in the definition of bleeding is likely responsible for the differences in estimates for the bleeding outcome and the composite outcome in the VALIDATE and target trial emulation.

Second, the observational estimates showed a risk difference of 1.0% (0.1%, 1.8%) for death after 14 days of follow-up (Figure 2, Appendix 6) whereas the randomized estimates found no discernible difference in mortality throughout the entire follow-up. This almost instantaneous difference in mortality suggests the presence of imbalanced, and unadjusted for, prognostic factors between treatment groups. It is possible, for example, that patients perceived to be at high risk of bleeding, and hence a higher immediate risk of death, may have been more likely to be administered bivalirudin (because bivalirudin was marketed as having a lower risk of bleeding than heparin). Even with rich data from the SWEDEHEART register, we were not able to accurately identify these potentially important characteristics. One way to aid future researchers in identifying important potential confounders would be if registers included detailed information on reasons for making treatment decisions.

The number of individuals randomized in VALIDATE was 6,006 (2,971 individuals either declined, were not asked, or could not be asked to participate), whereas the number of individuals included in the target trial emulation over a recruitment period of similar length was 4,940. This discrepancy is partly due to our inability to emulate the eligibility criteria for the VALIDATE trial. In the VALIDATE trial individuals given >5000U heparin before percutaneous coronary intervention were excluded, but the SWEDEHEART data does not include detail on dosage of heparin administered. We, therefore, excluded all 3,070 individuals given any dose of heparin before percutaneous coronary intervention, of whom some would have received low doses. A sensitivity analysis that did not exclude anyone given heparin before percutaneous coronary intervention (Appendix 12) included 7,652 individuals, and results for death and myocardial infarction by 180 days were broadly similar to the results from primary analyses. The risk difference for death between those given bivalirudin and heparin early in follow-up was, however, reduced in this sensitivity analysis, as shown in the inverse probability weighted Kaplan-Meier survival curves (Appendix 13). This suggests that by excluding those given any prior heparin in the main analysis, rather than only those given high dose heparin as in the trial, we may be selecting a group of individuals given heparin during percutaneous coronary intervention that are at lower risk of death soon after start of follow-up compared to those in the trial. Results from this sensitivity demonstrate the importance of designing a target trail that closely mimics the eligibility criteria of the index trial if aiming for comparable results, and is another possible explanation for the differences in short-term risk of death discussed above.

The SWEDEHEART register contains some of the most complete routinely collected data available worldwide when a patient undergoes percutaneous coronary intervention. Even with high quality data, however, close harmonization of protocols, adjustment for important confounders, and analytic methods appropriate for estimating causal quantities analogous to those estimated in the trial, we have shown that it is not always possible for the target trial emulation to obtain the same results as an index randomized trial due to trials collecting more detailed, study specific information that are not collected routinely. Nevertheless, we have shown that using real world data to emulate a target trial can deliver accurate long-term effect estimates in certain situations. It is, therefore, possible to foresee a system in which data from randomized trials with short-term follow-up are combined with real world data to estimate the long-term effects of certain interventions.^20^ Estimating the effect from the short randomized trial would overcome the problem of confounding soon after the intervention; we could then use observational data to estimate the effect during the remainder of follow-up and create synthetic survival curves using both data sources. This would enable prompt estimation of the long-term effects of interventions and allow adaptive clinical or regulatory changes to be put quickly into practice. Synergy between randomized trials and real-world data has the potential to revolutionize both the length of time patients can be followed up and the speed at which results from trials reach clinical practice.

## Data Availability

Pseudonymized personal data were obtained from national Swedish Registry holders after ethical approval and secrecy assessment. According to Swedish laws and regulations, personal sensitive data can only be made available for researchers who fulfil legal requirements for access to personal sensitive data.

## Footnotes

### Funding

This work was supported by grant from the Swedish Research Council (registration number 2018-03028). Issa J Dahabreh is supported by the Patient-Centered Outcomes Research Institute (PCORI, award ME-1502-27794)

### Disclosure

Dr. Matthews, Dr. Szummer, Dr. Lindahl, and Dr. Jernberg have nothing to disclose; Dr. Dahabreh reports grants from PCORI, during the conduct of the study; Dr. Erlinge reports personal fees from AstraZeneca, outside the submitted work; Dr. Feychting, Dr. Berglund, and Dr. Hernan report grants from Swedish Research Council, during the conduct of the study.

## Notes

### Author Declarations

Ethical approval was gained from the Swedish Ethical Review Authority

