## Appendix for "Comparing effect estimates in randomized trials and observational studies from the same population: an application to percutaneous coronary intervention"

| **Page** | **Appendix** |
| --- | --- |
| 2 | Appendix 1: Covariates definitions in the target trial |
| 4 | Appendix 2: Outcome definitions in the target trial |
| 5 | Appendix 3: Inverse probability weighted and standardization model specification |
| 7 | Appendix 4: Description of sensitivity analyses |
| 9 | Appendix 5: Unweighted survival curves from an observational emulation a target trial of bivalirudin vs. heparin, SWEDEHEART Register, 2012-2014 |
| 10 | Appendix 6: Estimated 14-day risk, risk difference, and risk ratios from the observational emulation of a target trial of bivalirudin vs. heparin, SWEDEHEART Register, 2012-2014, using inverse probability weighting models |
| 11 | Appendix 7: Estimated 30-day risk, risk difference, and risk ratios from the observational emulation of a target trial of bivalirudin vs. heparin, SWEDEHEART Register, 2012-2014, using inverse probability weighted models |
| 12 | Appendix 8: Estimated 180-day risk, risk difference, and risk ratios from the observational emulation of a target trial of bivalirudin vs. heparin, SWEDEHEART Register, 2012-2014, stratified by STEMI/NSTEMI, using inverse probability weighting models |
| 13 | Appendix 9: Sensitivity 1 - Estimated 180-day risk, risk difference, and risk ratios from the observational emulation of a target trial of bivalirudin vs. heparin, SWEDEHEART Register, 2012-2014, further excluding those with all cancer, using inverse probability weighting models |
| 14 | Appendix 10: Sensitivity 2 - Estimated 180-day risk, risk difference, and risk ratios from the observational emulation of a target trial of bivalirudin vs. heparin, SWEDEHEART Register, 2012-2014, excluding all patients over the age of 75 years, using inverse probability weighting models |
| 15 | Appendix 11: Sensitivity 3 - Estimated 180-day risk, risk difference, and risk ratios from the observational emulation of a target trial of bivalirudin vs. heparin, SWEDEHEART Register, 2012-2014, including only patients given P21Y2 inhibitors before percutaneous coronary intervention, using inverse probability weighting models |
| 16 | Appendix 12: Sensitivity 4 - Estimated 180-day risk, risk difference, and risk ratios from the observational emulation of a target trial of bivalirudin vs. heparin, SWEDEHEART Register, 2012-2014, including all individuals given heparin before percutaneous coronary intervention, using inverse probability weighting models |
| 17 | APPENDIX 13: Sensitivity 4 - Inverse-probability weighted survival curves from an observational emulation of a target trial of bivalirudin vs. heparin, SWEDEHEART Register, 2012-2014*, including all individuals given heparin before percutaneous coronary intervention |
| 18 | Appendix 14: Sensitivity 5 - Estimated 180-day risk, risk difference, and risk ratios from the observational emulation of a target trial of bivalirudin vs. heparin, SWEDEHEART Register, 2012-2014, including individuals missing GFR, and imputing median GFR and weight in analysis, using inverse probability weighting models |
| 19 | Appendix 15: Sensitivity 6 - Estimated 180-day risk, risk difference, and risk ratios from the observational emulation* of a target trial of bivalirudin vs. heparin, SWEDEHEART Register, 2012-2014, restricting to those that were given only bivalirudin or heparin, using inverse probability weighting models |
| 20 | Appendix 16: Sensitivity 7 - Estimated 180-day risk, risk difference, and risk ratios from the observational emulation of a target trial of bivalirudin vs. heparin, SWEDEHEART Register, 2012-2014, with expanded definition of the bleeding outcome, using inverse probability weighting models |
| 21 | Appendix 17: Sensitivity 8 - Estimated 180-day risk, risk difference, and risk ratios from the observational emulation of a target trial of bivalirudin vs. heparin, SWEDEHEART Register, 2012-2014, with 5 year look back period for confounder identified in the inpatient and outpatient register, using inverse probability weighting models |
| 22 | Appendix 18: Sensitivity 9 - Estimated 180-day risk, risk difference, and risk ratios from the observational emulation of a target trial of bivalirudin vs. heparin, SWEDEHEART Register, 2012-2014, using a complete case analysis, using inverse probability weighting models |
| 23 | Appendix 19: Sensitivity 10 - Estimated 180-day risk, risk difference, and risk ratios from the observational emulation of a target trial of bivalirudin vs. heparin, SWEDEHEART Register, 2012-2014, censoring at death, using inverse probability weighting models |

**APPENDICES**

**APPENDIX 1: Covariates definitions in the target trial**

| **Covariate** | **Register used** | **Definition (if further detail required)** | **Form** | **Categories** |
| --- | --- | --- | --- | --- |
| MI type | SCAAR |  | Indicator | STEMI/NSTEMI |
| Killip class | SCAAR |  | 5 categories | 1, 2, 3, 4, missing |
| Angiography finding | SCAAR | 0 – normal  1 – 1 vessel + no left main  2 – 2 vessels + no left main  3 – 3 vessels + no left main  4 – left main | 5 categories | 0, 1, 2, 3, 4 |
| Age | SCAAR |  | Linear, quadratic | N/A |
| Gender | SCAAR |  | Indicator | Male/female |
| Weight | SCAAR | Kilograms  No missing as to calculate GFR for eligibility, a non-missing weight is needed. | Linear, quadratic | N/A |
| Prior bleeding | Inpatient and outpatient registries | The patient registries were used to identify primary diagnoses of intracranial bleeding; major gastrointestinal bleeding; respiratory, renal/urinary tract, ocular, retroperitoneal or pericardial bleeding; and bleeding attributed to anemia within 3 years of PCI.  These outcomes were identified using the following ICD-10 codes: D62, I60, I61, I62, R31, R04, D500, H313, H356, H431, H450, I312, I850, K250, K252, K254, K256, K260, K262, K264, K266, K270, K272, K274, K276, K280, K282, K284, K286, K625, K661, K920, K921, K922, S064, S065, S066, J942, K228F, K298A, K638B, K638C, K838F, K868G, I864A, H052A, S368D, G951A | Indicator | Yes/No |
| Warfarin | SCAAR | Warfarin before or under PCI, if missing assumed no | Indicator | Yes/No |
| Anemia | RIKSHIA | Hemoglobin (g/L) was identified from the RIKSHIA register, then categorized to indicate severity of anemia Based on WHO anemia guidelines.  1 - Severe - lower than 70  2 - Moderate - 80-109  3 - Mild - 110-119 (women), 110-129 (men)  4 - No anemia - over 120 (women), over 130 (men)  9 - Missing | 5 categories | 1, 2, 3, 4, missing |
| GFR | SCAAR and RIKSHIA | First used creatinine recording from SCAAR register. If not available from SCAAR, then checked RIKSHIA register one month prior to PCI.  GFR then calculated using the Cockcroft-Gault formula: GFR = (((140-age)*weight)/(72*SCr)) (*0.85 if female)  No missing as eligibility criteria required to have GFR reading (excluded if severe renal disease or missing GFR) | Linear, quadratic | N/A |
| PCI Centre | SCAAR | Categorized into those that are part of a University Hospital: Karolinska Solna, Linköping, Örebro, Sahlgrenska, Lund, Malmö, Umeå, Uppsala. | Indicator | University hospital/Not university hospital |
| Prior myocardial infarction | SCAAR | If missing assumed no | Indicator | Yes/No |
| Prior percutaneous coronary intervention | SCAAR | If missing assumed no | Indicator | Yes/No |
| Prior coronary artery bypass grafting | SCAAR | If missing assumed no | Indicator | Yes/No |
| Diabetes | SCAAR | If missing assumed no | Indicator | Yes/No |
| Prior treatment for hypertension | SCAAR | If missing assumed no | Indicator | Yes/No |
| Prior lipid lowering treatment | SCAAR | If missing assumed no | Indicator | Yes/No |
| Smoking |  | 0 – never  1 – ex-smoker (> 1month)  2 – current  9 - missing | 4 categories | 0, 1, 2, missing |
| Heart rate | RIKSHIA | Some missing as captured from RIKSHIA and not all individuals have related RIKSHIA record  In main analysis missing imputed with median | Linear, quadratic | N/A |
| Systolic blood pressure | RIKSHIA | Some missing as captured from RIKSHIA and not all individuals have related RIKSHIA record  In main analysis missing imputed with median | Linear, quadratic | N/A |
| Diastolic blood pressure | RIKSHIA | Some missing as captured from RIKSHIA and not all individuals have related RIKSHIA record  In main analysis missing imputed with median | Linear, quadratic | N/A |

**APPENDIX 2: Outcome definitions in the target trial**

| **VALIDATE trial** | **Target trial definition** |
| --- | --- |
| Death from any cause | **1. SCAAR**  The SCAAR registry was used to identify date of death    **2. Cause of death register**  If there was no death record in the SCAAR register, but a record of death in the cause of death registry, then date of death was taken from cause of death register. |
| Myocardial infarction - the third universal myocardial definition (Thygesen et al Eur Heart J 2012;33:2551- 67) | **RIKSHIA**  **a. ≤ 2 days after PCI**  If there was a record of a myocardial infarction in the RIKSHIA registry within 2 days after PCI, then there also needed to be record of troponin levels greater than 40ng/L (both high sensitivity and conventional troponin, and I and T troponin) or a reinfarction. This was to ensure the record was not a repeat record of the original myocardial infarction.  **b. >2 days after PCI**  After 2 days, through until the end of follow up, all myocardial infarction records in the RIKSHIA registry were used (ICD 10 codes I21/I22). |
| Major bleeding event - adjudicated according to the BARC definition, type 2, 3, or 5 (Mehran R et al Circulation 2011; 123:2736-47) | **1. Inpatient and outpatient registries**  The patient registries were used to identify primary diagnoses of intracranial bleeding; major gastrointestinal bleeding; respiratory, renal/urinary tract, ocular, retroperitoneal or pericardial bleeding; and bleeding attributed to anemia.  These outcomes were identified using the following ICD-10 codes: D62, I60, I61, I62, R31, R04, D500, H313, H356, H431, H450, I312, I850, K250, K252, K254, K256, K260, K262, K264, K266, K270, K272, K274, K276, K280, K282, K284, K286, K625, K661, K920, K921, K922, S064, S065, S066, J942, K228F, K298A, K638B, K638C, K838F, K868G, I864A, H052A, S368D, G951A    **2. RIKSHIA**  The RIKSHIA registry also records if a patient bled within a period of care. This was used to identify lethal bleeding, cerebral bleeding, or bleeding that required transfusion. |
| Composite of all above | As above |

**APPENDIX 3: Inverse probability weighting and standardization**

In our target trial, the intention to treat effect is the effect of being assigned to bivalirudin versus heparin under percutaneous coronary intervention on the risk of death, myocardial infarction, bleeding, or a composite of all three. Estimating its observational analog requires adjustment for baseline confounders, which we did in two ways. First, we used inverse probability weighting of marginal structural models, and second, we used standardization.

**Inverse probability weighed models**

First, we estimate the stabilized inverse probability weight, *SW^A^*, truncated to the 99^th^ percentile, for each individual in the study population. Informally, the denominator of this weight is the probability that an individual received her observed treatment, *A,* given her measured confounders, *L*, and the numerator is the probability of receiving her observed treatment, *A*.

$$SW^{A}=f\left( A \right)/f\left( A | L \right)$$

Second, we fit pooled logistic regression models where individuals are weighted by their estimated *SW^A^*.

$$logit\left( Pr\left[ Y_{m+1}=0 | A,Y_{m}=0 \right] \right)=\alpha_{0,m}+\alpha_{1}A+\alpha_{2}Am+\alpha_{3}Am^{2}$$

where:
*Y_m+1_* = Indicator for the outcome of interest at time *m+1*.
*A* = Indicator for the treatment group.
$\alpha_{0,m}$ = Time-varying intercept estimated as a constant plus linear and quadratic terms at time *m*.

The predicted probabilities from the inverse probability weighted hazards model can then be multiplied over time, in the absence of censoring, to obtain an estimate of survival under each treatment strategy at each time, *m*.

$$\hat{{IPW}_{k}^{a}}=\prod_{m=1}^{k} expit\left( \hat{\alpha}_{0,m}+\hat{\alpha}_{1}a+\hat{\alpha}_{2}am+\hat{\alpha}_{3}am^{2} \right)$$

The risk at time, *k*, can then be estimated by taking one minus $\hat{{IPW}_{k}^{a}}$, and risk differences and risk ratios can be calculated. Finally, we use nonparametric bootstrapping with 500 samples to calculate all 95% confidence intervals.

**Standardization models**

First, we fit the pooled logistic regression model.

$$logit\left( Pr\left[ Y_{m+1}=0 | A,{L, Y}_{m}=0 \right] \right)=\beta_{0,m}+\beta_{1}A+\beta_{2}Am+\beta_{3}Am^{2}+\beta_{4}^{T}L$$

where:
*Y_m+1_* = Indicator for the outcome of interest at time *m+1*.
*A* = Indicator for the treatment group.
*L* = Vector of potential confounders at baseline
$\beta_{0,m}$ = Time-varying intercept estimated as a constant plus linear and quadratic terms at time *m*.

We then use the predicted probabilities from these models, multiplied over time, in the absence of censoring, to obtain an estimate of survival of each individual, *i*, under each treatment strategy at each time, *m*, conditional on the individual’s baseline confounders, *L_i_.*

$$\hat{{OM}_{\text{i},k}^{a}}=\prod_{m=1}^{k} expit\left( \hat{\beta}_{0,m}+\hat{\beta}_{1}a+\hat{\beta}_{2}am+\hat{\beta}_{3}am^{2}+\hat{\beta}_{4}^{T}L_{i} \right)$$

We then standardized the survival probabilities at each time point to the empirical distribution of the confounders at baseline.

$$\hat{{OM}_{k}^{a}}=\frac{1}{n}\sum_{i=1}^{n} \hat{{OM}_{\text{i},k}^{a}}$$

Where *n* is the number of individuals in the study population. The risk at time, *k*, can then be estimated by taking one minus $\hat{{OM}_{k}^{a}}$, and risk differences and risk ratios can be calculated. Finally, we used nonparametric bootstrapping with 500 samples to calculate all 95% confidence intervals.

**APPENDIX 4: Description of sensitivity analyses**

**Sensitivity analysis 1: eligibility criteria 1**

We expanded the definition of diagnoses that could indicate a life expectancy of less than a year (one of the exclusion criteria) to include all cancer diagnoses (ICD-10 code: C).

**Sensitivity analysis 2: eligibility criteria 2**

We excluded all patients over the age of 75 years at the time of PCI.

**Sensitivity analysis 3: eligibility criteria 3**

We only included those who were recorded as given P21Y2 before percutaneous coronary intervention, not under percutaneous intervention

**Sensitivity analysis 4: eligibility criteria 4**

We included everyone given any dose of heparin before coronary intervention

**Sensitivity analysis 5: eligibility criteria 5**

We included everyone with a missing GFR and imputed missing GFR and weight variables with the median GFR and weight in the analysis.

**Sensitivity analysis 6: treatment strategy**

We restricted treatment to those who were only given bivalirudin or heparin (original definition is exposure to bivalirudin if given both treatments as assumed that heparin is low dose if have been given both). 1,466 of the 1,951 (75%) of the patients originally assigned to bivalirudin were also given low dose heparin at the same time. These patients were excluded from this analysis.

**Sensitivity analysis 7: outcome**

We expanded the definition of bleeding to include less severe bleeding outcomes. The additional ICD-10 codes we identified in the inpatient and outpatient registers were:

- anal K62.5
- anovulatory N97.0
- atonic, following delivery O72.1
- capillary I78.8
  - puerperal O72.2
- contact N93.0
- due to uterine subinvolution N85.3
- ear H92.2
- excessive, associated with menopausal onset N92.4
- following intercourse N93.0
- gastrointestinal K92.2
- hemorrhoids K64
- intermenstrual (regular) N92.3
  - irregular N92.1
- irregular N92.6
- menopausal N92.4
- nipple N64.59
- nose R04.0
- ovulation N92.3
- perimenopausal N92.4
- postclimacteric N95.0
- postcoital N93.0
- postmenopausal N95.0
- pre-pubertal vaginal N93.1
- preclimacteric N92.4
- puberty N92.2 (excessive, with onset of menstrual periods)
- rectum, rectal K62.5
  - newborn P54.2
- throat R04.1
- tooth socket K91.840 (post-extraction)
- umbilical stump P51.9
- uterus, uterine NEC N93.9
  - climacteric N92.4
  - dysfunctional or functional N93.8
  - menopausal N92.4
  - preclimacteric or premenopausal N92.4
  - unrelated to menstrual cycle N93.9
- vagina, vaginal (abnormal) N93.9
  - dysfunctional or functional N93.8
  - newborn P54.6
  - pre-pubertal N93.1
- vicarious N94.89

**Sensitivity analysis 8: confounders**

We changed the definition to the confounders that were defined in the inpatient, outpatient, and prescribed drug registers (bleeding, warfarin, and NOAC), so they were identified from a 5-year look back period rather than 3-year.

**Sensitivity analysis 9: missing data**

We carried out a complete case analysis, excluding patients that had missing data for the Killip class and anemia variables (18%).

**Sensitivity analysis 10: censoring at death**

In the analyses for the myocardial infarction and bleeding outcomes, we censored individuals at date of death.

**APPENDIX 5: Unweighted survival curves from an observational emulation a target trial of bivalirudin vs. heparin, SWEDEHEART Register, 2012-2014**

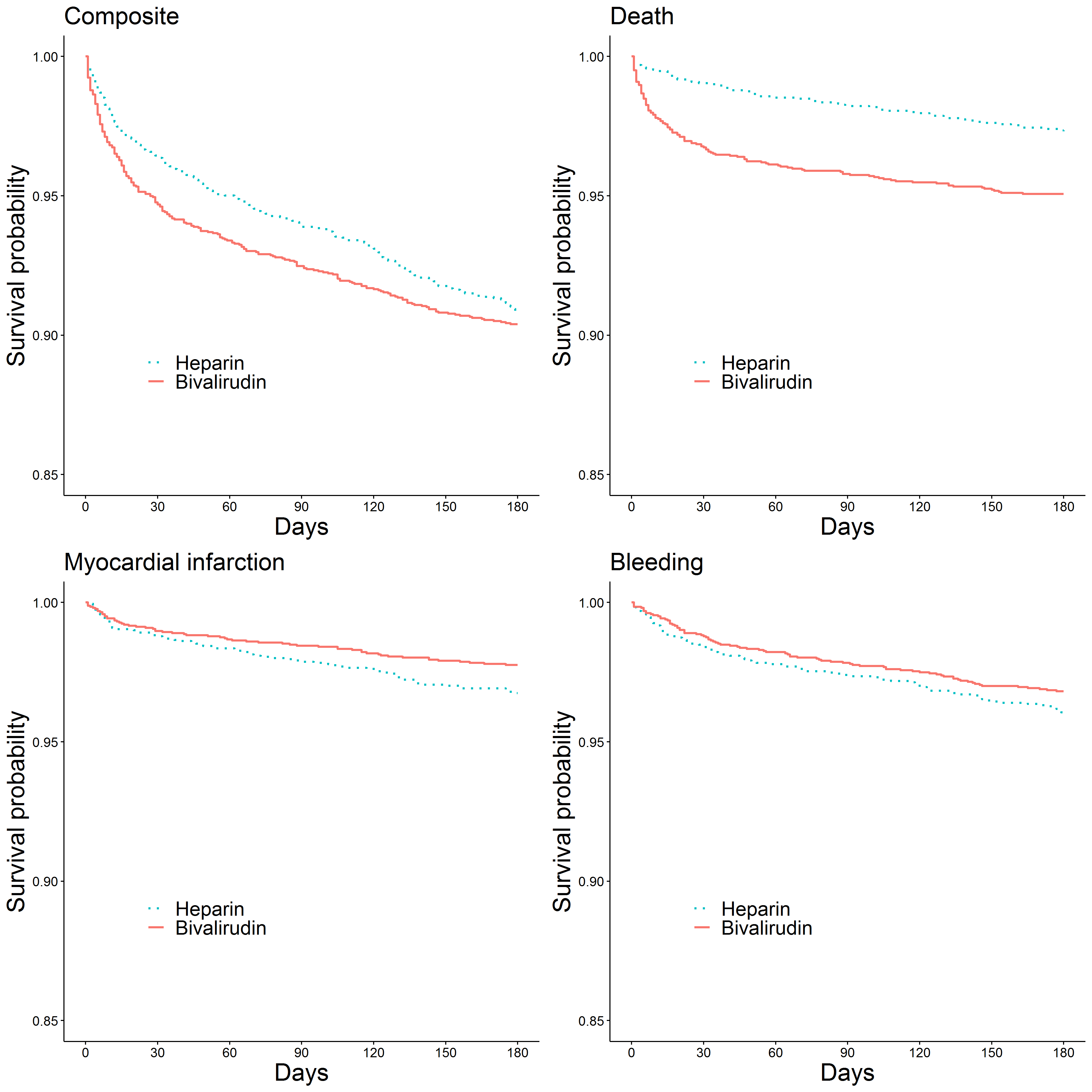

| **APPENDIX 6: Estimated 14-day risk, risk difference, and risk ratios from the observational emulation* of a target trial of bivalirudin vs. heparin, SWEDEHEART Register, 2012-2014, using inverse probability weighting models** | | | | |
| --- | --- | --- | --- | --- |
| **Outcome** | **Risk (%, 95% CI)** | | **Risk difference (95% CI)** | **Risk ratio (95% CI)** |
|  | Bivalirudin | Heparin |  |  |
| Composite | 3.7 (2.9,4.4) | 3.3 (2.4,4.1) | 0.4 (-0.7,1.5) | 1.12 (0.79,1.58) |
| Death | 2.1 (1.5,2.6) | 1.1 (0.5,1.7) | 1.0 (0.1,1.8) | 1.85 (0.95,3.63) |
| Myocardial infarction | 1.1 (0.6,1.6) | 0.9 (0.6,1.3) | 0.2 (-0.5,0.8) | 1.16 (0.58,2.32) |
| Bleeding | 0.6 (0.3,0.9) | 1.3 (0.8,1.8) | -0.7 (-1.3,-0.1) | 0.46 (0.22,0.98) |
| * Adjusted at baseline for severity of myocardial infarction (STEMI/NSTEMI, Killip class, angiography finding), center where percutaneous coronary intervention took place (university hospital/non-university hospital), demographics (sex, age), lifestyle characteristics (weight, smoking), laboratory measurements (GFR, heart rate, systolic blood pressure, diastolic blood pressure), diagnoses (diabetes, severe bleeding, anemia), medications (warfarin, anti-hypertensives, lipid lowering treatment), and prior cardiovascular disease or cardiovascular procedures (myocardial infarction, percutaneous coronary intervention, coronary artery bypass grafting) | | | | |

| **APPENDIX 7: Estimated 30-day risk, risk difference, and risk ratios from the observational emulation* of a target trial of bivalirudin vs. heparin, SWEDEHEART Register, 2012-2014, using inverse probability weighting models** | | | | |
| --- | --- | --- | --- | --- |
| **Outcome** | **Risk (%, 95% CI)** | | **Risk difference (95% CI)** | **Risk ratio (95% CI)** |
|  | Bivalirudin | Heparin |  |  |
| Composite | 5.1 (4.3,6.0) | 4.6 (3.6,5.66) | 0.5 (-0.8,1.8) | 1.10 (0.83,1.46) |
| Death | 2.7 (2.1,3.3) | 1.7 (1.0,2.39) | 1.0 (0.1,1.9) | 1.61 (0.99,2.63) |
| Myocardial infarction | 1.4 (0.8,2.0) | 1.1 (0.7,1.56) | 0.3 (-0.4,1.0) | 1.22 (0.68,2.19) |
| Bleeding | 1.3 (0.8,1.7) | 2.0 (1.4,2.69) | -0.8 (-1.6,0.1) | 0.63 (0.37,1.06) |
| * Adjusted at baseline for severity of myocardial infarction (STEMI/NSTEMI, Killip class, angiography finding), center where percutaneous coronary intervention took place (university hospital/non-university hospital), demographics (sex, age), lifestyle characteristics (weight, smoking), laboratory measurements (GFR, heart rate, systolic blood pressure, diastolic blood pressure), diagnoses (diabetes, severe bleeding, anemia), medications (warfarin, anti-hypertensives, lipid lowering treatment), and prior cardiovascular disease or cardiovascular procedures (myocardial infarction, percutaneous coronary intervention, coronary artery bypass grafting) | | | | |

| **APPENDIX 8: Estimated 180-day risk, risk difference, and risk ratios from the observational emulation* of a target trial of bivalirudin vs. heparin, SWEDEHEART Register, 2012-2014, stratified by STEMI/NSTEMI , using inverse probability weighting models** | | | | |
| --- | --- | --- | --- | --- |
| **Outcome** | **Risk (%, 95% CI)** | | **Risk difference (95% CI)** | **Risk ratio (95% CI)** |
|  | Bivalirudin | Heparin |  |  |
| ***STEMI*** |  |  |  |  |
| Composite | 10.4 (9.3,11.4) | 11.9 (9.3,14.6) | -1.6 (-4.4,1.2) | 0.87 (0.68,1.11) |
| Death | 5.5 (4.8,6.3) | 4.3 (2.6,6.1) | 1.2 (-0.7,3.1) | 1.28 (0.79,2.07) |
| Myocardial infarction | 2.1 (1.6,2.5) | 2.5 (1.5,3.6) | -0.5 (-1.6,0.7) | 0.81 (0.48,1.37) |
| Bleeding | 3.4 (2.8,4.0) | 5.9 (3.9,7.9) | -2.6 (-4.6,-0.5) | 0.57 (0.39,0.84) |
| ***NSTEMI*** |  |  |  |  |
| Composite | 8.4 (6.4,10.5) | 8.5 (7.5,9.4) | 0.0 (-2.3,2.3) | 1.00 (0.75,1.33) |
| Death | 2.7 (1.6,3.7) | 2.4 (1.9,2.9) | 0.3 (-0.9,1.5) | 1.11 (0.68,1.82) |
| Myocardial infarction | 4.1 (2.6,5.7) | 3.0 (2.5,3.6) | 1.1 (-0.6,2.7) | 1.36 (0.86,2.15) |
| Bleeding | 3.1 (1.9,4.4) | 3.5 (2.9,4.2) | -0.4 (-1.8,1.0) | 0.89 (0.54,1.46) |
| * Adjusted at baseline for severity of myocardial infarction (STEMI/NSTEMI, Killip class, angiography finding), center where percutaneous coronary intervention took place (university hospital/non-university hospital), demographics (sex, age), lifestyle characteristics (weight, smoking), laboratory measurements (GFR, heart rate, systolic blood pressure, diastolic blood pressure), diagnoses (diabetes, severe bleeding, anemia), medications (warfarin, anti-hypertensives, lipid lowering treatment), and prior cardiovascular disease or cardiovascular procedures (myocardial infarction, percutaneous coronary intervention, coronary artery bypass grafting) | | | | |

| **APPENDIX 9: Sensitivity 1 - Estimated 180-day risk, risk difference, and risk ratios from the observational emulation* of a target trial of bivalirudin vs. heparin, SWEDEHEART Register, 2012-2014, further excluding those with all cancer, using inverse probability weighting models (N=4,527)** | | | | |
| --- | --- | --- | --- | --- |
| **Outcome** | **Risk (%, 95% CI)** | | **Risk difference (95% CI)** | **Risk ratio (95% CI)** |
|  | Bivalirudin | Heparin |  |  |
| Composite | 9.0 (7.9,10.1) | 9.0 (7.7,10.3) | -0.1 (-1.8,1.7) | 0.99 (0.81,1.21) |
| Death | 3.8 (3.1,4.5) | 2.9 (2.0,3.8) | 0.9 (-0.2,2.0) | 1.30 (0.90,1.87) |
| Myocardial infarction | 3.1 (2.3,3.9) | 2.6 (2.0,3.2) | 0.5 (-0.5,1.6) | 1.20 (0.81,1.76) |
| Bleeding | 3.1 (2.4,3.7) | 4.3 (3.4,5.3) | -1.3 (-2.4,-0.1) | 0.71 (0.51,0.98) |
| * Adjusted at baseline for severity of myocardial infarction (STEMI/NSTEMI, Killip class, angiography finding), center where percutaneous coronary intervention took place (university hospital/non-university hospital), demographics (sex, age), lifestyle characteristics (weight, smoking), laboratory measurements (GFR, heart rate, systolic blood pressure, diastolic blood pressure), diagnoses (diabetes, severe bleeding, anemia), medications (warfarin, anti-hypertensives, lipid lowering treatment), and prior cardiovascular disease or cardiovascular procedures (myocardial infarction, percutaneous coronary intervention, coronary artery bypass grafting) | | | | |

| **APPENDIX 10: Sensitivity 2 - Estimated 180-day risk, risk difference, and risk ratios from the observational emulation* of a target trial of bivalirudin vs. heparin, SWEDEHEART Register, 2012-2014, excluding all patients over the age of 75 years, using inverse probability weighting models (N=3,624)** | | | | |
| --- | --- | --- | --- | --- |
| **Outcome** | **Risk (%, 95% CI)** | | **Risk difference (95% CI)** | **Risk ratio (95% CI)** |
|  | Bivalirudin | Heparin |  |  |
| Composite | 6.7 (5.5,7.9) | 6.7 (5.3,8.0) | 0.0 (-1.7,1.7) | 1.00 (0.77,1.31) |
| Death | 2.4 (1.7,3.1) | 1.5 (0.9,2.1) | 0.9 (0.0,1.8) | 1.57 (0.91,2.69) |
| Myocardial infarction | 2.5 (1.6,3.4) | 2.0 (1.4,2.5) | 0.5 (-0.5,1.6) | 1.28 (0.78,2.08) |
| Bleeding | 2.6 (2.0,3.3) | 3.6 (2.6,4.6) | -1.0 (-2.1,0.2) | 0.73 (0.50,1.07) |
| * Adjusted at baseline for severity of myocardial infarction (STEMI/NSTEMI, Killip class, angiography finding), center where percutaneous coronary intervention took place (university hospital/non-university hospital), demographics (sex, age), lifestyle characteristics (weight, smoking), laboratory measurements (GFR, heart rate, systolic blood pressure, diastolic blood pressure), diagnoses (diabetes, severe bleeding, anemia), medications (warfarin, anti-hypertensives, lipid lowering treatment), and prior cardiovascular disease or cardiovascular procedures (myocardial infarction, percutaneous coronary intervention, coronary artery bypass grafting) | | | | |

| **APPENDIX 11: Sensitivity 3 - Estimated 180-day risk, risk difference, and risk ratios from the observational emulation* of a target trial of bivalirudin vs. heparin, SWEDEHEART Register, 2012-2014, including only patients given P21Y2 inhibitors before percutaneous coronary intervention, using inverse probability weighting models (N=4,113)** | | | | |
| --- | --- | --- | --- | --- |
| **Outcome** | **Risk (%, 95% CI)** | | **Risk difference (95% CI)** | **Risk ratio (95% CI)** |
|  | Bivalirudin | Heparin |  |  |
| Composite | 9.0 (7.7,10.3) | 9.5 (8.3,10.7) | -0.5 (-2.2,1.3) | 0.95 (0.78,1.16) |
| Death | 3.5 (2.8,4.3) | 3.1 (2.3,3.9) | 0.5 (-0.6,1.5) | 1.15 (0.82,1.63) |
| Myocardial infarction | 3.5 (2.5,4.5) | 3.0 (2.4,3.6) | 0.5 (-0.7,1.6) | 1.16 (0.80,1.69) |
| Bleeding | 3.1 (2.4,3.9) | 4.3 (3.4,5.3) | -1.2 (-2.4,0.0) | 0.73 (0.52,1.02) |
| * Adjusted at baseline for severity of myocardial infarction (STEMI/NSTEMI, Killip class, angiography finding), center where percutaneous coronary intervention took place (university hospital/non-university hospital), demographics (sex, age), lifestyle characteristics (weight, smoking), laboratory measurements (GFR, heart rate, systolic blood pressure, diastolic blood pressure), diagnoses (diabetes, severe bleeding, anemia), medications (warfarin, anti-hypertensives, lipid lowering treatment), and prior cardiovascular disease or cardiovascular procedures (myocardial infarction, percutaneous coronary intervention, coronary artery bypass grafting) | | | | |

| **APPENDIX 12: Sensitivity 4 - Estimated 180-day risk, risk difference, and risk ratios from the observational emulation* of a target trial of bivalirudin vs. heparin, SWEDEHEART Register, 2012-2014, including all individuals given heparin before percutaneous coronary intervention, using inverse probability weighting models (N=7,652)** | | | | |
| --- | --- | --- | --- | --- |
| **Outcome** | **Risk (%, 95% CI)** | | **Risk difference (95% CI)** | **Risk ratio (95% CI)** |
|  | Bivalirudin | Heparin |  |  |
| Composite | 9.4 (8.6,10.2) | 10.0 (8.7,11.2) | -0.5 (-2.0,0.9) | 0.94 (0.81,1.10) |
| Death | 3.9 (3.5,4.4) | 3.6 (2.7,4.4) | 0.4 (-0.5,1.3) | 1.11 (0.86,1.44) |
| Myocardial infarction | 2.6 (2.1,3.1) | 2.6 (2.1,3.2) | 0.0 (-0.8,0.7) | 0.99 (0.73,1.34) |
| Bleeding | 3.7 (3.3,4.2) | 4.4 (3.5,5.2) | -0.6 (-1.6,0.3) | 0.86 (0.67,1.09) |
| * Adjusted at baseline for severity of myocardial infarction (STEMI/NSTEMI, Killip class, angiography finding), center where percutaneous coronary intervention took place (university hospital/non-university hospital), demographics (sex, age), lifestyle characteristics (weight, smoking), laboratory measurements (GFR, heart rate, systolic blood pressure, diastolic blood pressure), diagnoses (diabetes, severe bleeding, anemia), medications (warfarin, anti-hypertensives, lipid lowering treatment), and prior cardiovascular disease or cardiovascular procedures (myocardial infarction, percutaneous coronary intervention, coronary artery bypass grafting) | | | | |

**APPENDIX 13: Sensitivity 4 - Inverse-probability weighted survival curves from an observational emulation of a target trial of bivalirudin vs. heparin, SWEDEHEART Register, 2012-2014*, including all individuals given heparin before percutaneous coronary intervention**
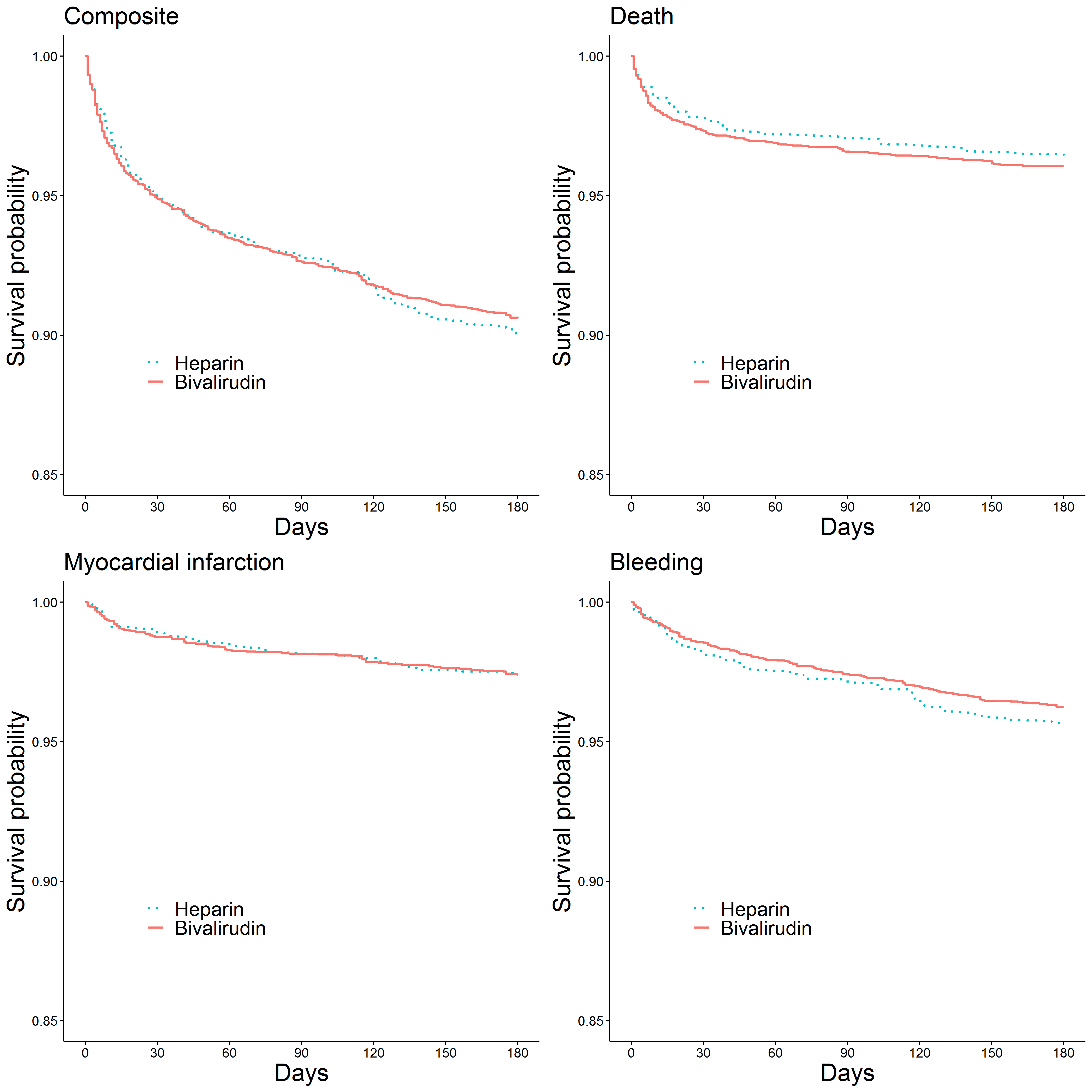
**(N=7,652)**

*Adjusted at baseline for severity of myocardial infarction (STEMI/NSTEMI, Killip class, angiography finding), center where percutaneous coronary intervention took place (university hospital/non-university hospital), demographics (sex, age), lifestyle characteristics (weight, smoking), laboratory measurements (GFR, heart rate, systolic blood pressure, diastolic blood pressure), diagnoses (diabetes, severe bleeding, anemia), medications (warfarin, anti-hypertensives, lipid lowering treatment), and prior cardiovascular disease or cardiovascular procedures (myocardial infarction, percutaneous coronary intervention, coronary artery bypass grafting)

| **APPENDIX 14: Sensitivity 5 - Estimated 180-day risk, risk difference, and risk ratios from the observational emulation* of a target trial of bivalirudin vs. heparin, SWEDEHEART Register, 2012-2014, including individuals missing GFR, and imputing median GFR and weight, using inverse probability weighting models (N=6,172)** | | | | |
| --- | --- | --- | --- | --- |
| **Outcome** | **Risk (%, 95% CI)** | | **Risk difference (95% CI)** | **Risk ratio (95% CI)** |
|  | Bivalirudin | Heparin |  |  |
| Composite | 9.3 (8.4,10.1) | 9.4 (8.2,10.6) | -0.2 (-1.7,1.3) | 0.98 (0.83,1.16) |
| Death | 4.2 (3.6,4.8) | 3.6 (2.7,4.4) | 0.6 (-0.4,1.7) | 1.18 (0.89,1.57) |
| Myocardial infarction | 2.8 (2.2,3.5) | 2.5 (2.0,3.0) | 0.3 (-0.4,1.1) | 1.14 (0.84,1.55) |
| Bleeding | 3.1 (2.6,3.6) | 4.0 (3.2,4.8) | -0.9 (-1.9,0.0) | 0.77 (0.58,1.01) |
| * Adjusted at baseline for severity of myocardial infarction (STEMI/NSTEMI, Killip class, angiography finding), center where percutaneous coronary intervention took place (university hospital/non-university hospital), demographics (sex, age), lifestyle characteristics (weight, smoking), laboratory measurements (GFR, heart rate, systolic blood pressure, diastolic blood pressure), diagnoses (diabetes, severe bleeding, anemia), medications (warfarin, anti-hypertensives, lipid lowering treatment), and prior cardiovascular disease or cardiovascular procedures (myocardial infarction, percutaneous coronary intervention, coronary artery bypass grafting) | | | | |

| **APPENDIX 15: Sensitivity 6 - Estimated 180-day risk, risk difference, and risk ratios from the observational emulation* of a target trial of bivalirudin vs. heparin, SWEDEHEART Register, 2012-2014, restricting to those that were given only bivalirudin or heparin, using inverse probability weighting models (N=2,886)** | | | | |
| --- | --- | --- | --- | --- |
| **Outcome** | **Risk (%, 95% CI)** | | **Risk difference (95% CI)** | **Risk ratio (95% CI)** |
|  | Bivalirudin | Heparin |  |  |
| Composite | 7.3 (4.7,9.9) | 9.7 (8.7,10.8) | -2.4 (-5.2, 0.4) | 0.75 (0.51,1.12) |
| Death | 4.4 (2.1,6.7) | 3.0 (2.4,3.7) | 1.4 (-1.1, 3.8) | 1.45 (0.76,2.77) |
| Myocardial infarction | 1.6 (0.5,2.7) | 3.1 (2.5,3.6) | -1.5 (-2.6,-0.3) | 0.52 (0.22,1.23) |
| Bleeding | 2.6 (0.9,4.3) | 4.4 (3.6,5.1) | -1.7 (-3.6, 0.1) | 0.60 (0.27,1.34) |
| * Adjusted at baseline for severity of myocardial infarction (STEMI/NSTEMI, Killip class, angiography finding), center where percutaneous coronary intervention took place (university hospital/non-university hospital), demographics (sex, age), lifestyle characteristics (weight, smoking), laboratory measurements (GFR, heart rate, systolic blood pressure, diastolic blood pressure), diagnoses (diabetes, severe bleeding, anemia), medications (warfarin, anti-hypertensives, lipid lowering treatment), and prior cardiovascular disease or cardiovascular procedures (myocardial infarction, percutaneous coronary intervention, coronary artery bypass grafting) | | | | |

| **APPENDIX 16: Sensitivity 7 - Estimated 180-day risk, risk difference, and risk ratios from the observational emulation* of a target trial of bivalirudin vs. heparin, SWEDEHEART Register, 2012-2014, with expanded definition of the bleeding outcome, using inverse probability weighting models (N=4,940)** | | | | |
| --- | --- | --- | --- | --- |
| **Outcome** | **Risk (%, 95% CI)** | | **Risk difference (95% CI)** | **Risk ratio (95% CI)** |
|  | Bivalirudin | Heparin |  |  |
| Composite | 9.5 (8.4,10.6) | 10.2 (8.8,11.5) | -0.7 (-2.4,1.1) | 0.94 (0.78,1.12) |
| Bleeding | 3.5 (2.8,4.2) | 4.8 (3.8,5.7) | -1.3 (-2.5,-0.1) | 0.74 (0.54,0.99) |
| * Adjusted at baseline for severity of myocardial infarction (STEMI/NSTEMI, Killip class, angiography finding), center where percutaneous coronary intervention took place (university hospital/non-university hospital), demographics (sex, age), lifestyle characteristics (weight, smoking), laboratory measurements (GFR, heart rate, systolic blood pressure, diastolic blood pressure), diagnoses (diabetes, severe bleeding, anemia), medications (warfarin, anti-hypertensives, lipid lowering treatment), and prior cardiovascular disease or cardiovascular procedures (myocardial infarction, percutaneous coronary intervention, coronary artery bypass grafting) | | | | |

| **APPENDIX 17: Sensitivity 8 - Estimated 180-day risk, risk difference, and risk ratios from the observational emulation* of a target trial of bivalirudin vs. heparin, SWEDEHEART Register, 2012-2014, with 5 year look back period for confounder identified in the inpatient and outpatient register, using inverse probability weighting models (N=4,940)** | | | | |
| --- | --- | --- | --- | --- |
| **Outcome** | **Risk (%, 95% CI)** | | **Risk difference (95% CI)** | **Risk ratio (95% CI)** |
|  | Bivalirudin | Heparin |  |  |
| Composite | 9.3 (8.2,10.4) | 10.0 (8.7,11.3) | -0.7 (-2.5,1.0) | 0.93 (0.77,1.12) |
| Death | 4.1 (3.4,4.8) | 3.4 (2.5,4.2) | 0.7 (-0.4,1.8) | 1.21 (0.89,1.66) |
| Myocardial infarction | 3.0 (2.3,3.8) | 2.8 (2.2,3.4) | 0.2 (-0.7,1.2) | 1.08 (0.77,1.52) |
| Bleeding | 3.2 (2.5,3.8) | 4.6 (3.7,5.6) | -1.4 (-2.6,-0.3) | 0.69 (0.50,0.94) |
| * Adjusted at baseline for severity of myocardial infarction (STEMI/NSTEMI, Killip class, angiography finding), center where percutaneous coronary intervention took place (university hospital/non-university hospital), demographics (sex, age), lifestyle characteristics (weight, smoking), laboratory measurements (GFR, heart rate, systolic blood pressure, diastolic blood pressure), diagnoses (diabetes, severe bleeding, anemia), medications (warfarin, anti-hypertensives, lipid lowering treatment), and prior cardiovascular disease or cardiovascular procedures (myocardial infarction, percutaneous coronary intervention, coronary artery bypass grafting) | | | | |

| **APPENDIX 18: Sensitivity 9 - Estimated 180-day risk, risk difference, and risk ratios from the observational emulation* of a target trial of bivalirudin vs. heparin, SWEDEHEART Register, 2012-2014, using a complete case analysis, using inverse probability weighting models (N=3,704)** | | | | |
| --- | --- | --- | --- | --- |
| **Outcome** | **Risk (%, 95% CI)** | | **Risk difference (95% CI)** | **Risk ratio (95% CI)** |
|  | Bivalirudin | Heparin |  |  |
| Composite | 9.2 (8.0, 10.4) | 9.2 (7.9, 10.6) | 0.0 (-1.9, 1.8) | 1.00 (0.81, 1.22) |
| Death | 3.6 (2.9, 4.3) | 2.4 (1.6, 3.2) | 1.2 (0.1, 2.2) | 1.49 (1.00, 2.23) |
| Myocardial infarction | 3.1 (2.3, 4.0) | 2.4 (1.8, 3.0) | 0.7 (-0.4, 1.8) | 1.29 (0.85, 1.96) |
| Bleeding | 3.3 (2.6, 4.1) | 4.8 (3.6, 6.0) | -1.5 (-2.9, 0.0) | 0.70 (0.49, 1.00) |
| * Adjusted at baseline for severity of myocardial infarction (STEMI/NSTEMI, Killip class, angiography finding), center where percutaneous coronary intervention took place (university hospital/non-university hospital), demographics (sex, age), lifestyle characteristics (weight, smoking), laboratory measurements (GFR, heart rate, systolic blood pressure, diastolic blood pressure), diagnoses (diabetes, severe bleeding, anemia), medications (warfarin, anti-hypertensives, lipid lowering treatment), and prior cardiovascular disease or cardiovascular procedures (myocardial infarction, percutaneous coronary intervention, coronary artery bypass grafting) | | | | |

| **APPENDIX 19: Sensitivity 10 - Estimated 180-day risk, risk difference, and risk ratios from the observational emulation* of a target trial of bivalirudin vs. heparin, SWEDEHEART Register, 2012-2014, censoring at death, using inverse probability weighting models (N=4,940)** | | | | |
| --- | --- | --- | --- | --- |
| **Outcome** | **Risk (%, 95% CI)** | | **Risk difference (95% CI)** | **Risk ratio (95% CI)** |
|  | Bivalirudin | Heparin |  |  |
| Myocardial infarction | 3.1 (2.3, 3.9) | 2.9 (2.2, 3.5) | 0.3 (-0.8, 1.3) | 1.09 (0.76, 1.55) |
| Bleeding | 3.3 (2.6, 4.0) | 4.7 (3.7, 5.7) | -1.4 (-2.7, -0.2) | 0.69 (0.51, 0.95) |
| * Adjusted at baseline for severity of myocardial infarction (STEMI/NSTEMI, Killip class, angiography finding), center where percutaneous coronary intervention took place (university hospital/non-university hospital), demographics (sex, age), lifestyle characteristics (weight, smoking), laboratory measurements (GFR, heart rate, systolic blood pressure, diastolic blood pressure), diagnoses (diabetes, severe bleeding, anemia), medications (warfarin, anti-hypertensives, lipid lowering treatment), and prior cardiovascular disease or cardiovascular procedures (myocardial infarction, percutaneous coronary intervention, coronary artery bypass grafting) | | | | |
